# Effect of household use of multiple micronutrient-fortified bouillon on micronutrient status among women and children in two districts in the Northern Region of Ghana: Protocol for the Condiment Micronutrient Innovation Trial (CoMIT), a community-based randomized controlled trial

**DOI:** 10.1101/2023.07.19.23292899

**Authors:** Reina Engle-Stone, K Ryan Wessells, Marjorie J. Haskell, Sika M. Kumordzie, Charles D. Arnold, Jennie N. Davis, Emily R. Becher, Ahmed D. Fuseini, Kania W. Nyaaba, Xiuping (Jenny) Tan, Katherine P. Adams, Georg Lietz, Stephen A. Vosti, Seth Adu-Afarwuah

**Author notes:** Corresponding author: (RES).

## Abstract

**Introduction:** Micronutrient deficiencies are prevalent in West Africa, particularly among women of reproductive age (WRA) and young children. Bouillon is a promising food fortification vehicle due to its widespread consumption. This study aims to evaluate the impact of multiple micronutrient-fortified bouillon cubes, compared to control bouillon cubes (fortified with iodine only), on micronutrient status and hemoglobin concentrations among lactating and non-lactating WRA and young children in northern Ghana.

**Methods:** This randomized, controlled doubly-masked trial will be conducted in the Kumbungu and Tolon districts in the Northern Region of Ghana, where prior data indicate multiple micronutrient deficiencies are common. Participants will be: 1) non-pregnant non-lactating WRA (15-49 y), 2) children 2-5 y, and 3) non-pregnant lactating women 4-18 months postpartum. Eligible participants will be randomly assigned to receive household rations of one of two types of bouillon cubes: 1) a multiple micronutrient-fortified bouillon cube containing vitamin A, folic acid, vitamin B12, iron, zinc, and iodine, or 2) a control cube containing iodine only.

Each participant’s household will receive a ration of bouillon cubes every 2 weeks, and households will be advised to prepare meals as usual, using the study-provided cubes. The trial duration will be 9 months for non-pregnant non-lactating WRA and children, and 3 months for lactating women. The primary outcomes will be changes in biomarkers of micronutrient status and hemoglobin. Secondary outcomes will include change in prevalence of micronutrient deficiency and anemia; dietary intake of bouillon and micronutrients; inflammation, malaria, and morbidity symptoms; and child growth and development.

**Discussion:** Evidence from this study will inform discussions about bouillon fortification in Ghana and West Africa.

## Introduction

Micronutrient deficiencies are severe and widespread in West Africa, particularly among young children, pregnant and lactating women, and women of reproductive age [1–3]. These deficiencies have developmental and physical consequences, including increased risk of morbidity and mortality [4], which, in turn, have economic consequences [5]. While various programs to address micronutrient deficiencies have been scaled up, including industrial fortification of cooking oil and wheat flour with micronutrients [6], it is unlikely that a single such program will eliminate all micronutrient deficiencies, since delivery platforms may provide only a subset of nutrients or reach only a subset of the deficient population.

In Ghana, micronutrient deficiencies remain common, particularly among women and children, despite the presence of various decades-long micronutrient intervention programs [7]. For example, in the 2017 Ghana Micronutrient Survey, the percentage of preschool children who had iron deficiency (serum ferritin <12 μg/L adjusted for inflammation) was 40% for the Northern Belt, 18% for the Middle Belt, and 13% for the Southern Belt [8]. Likewise, the percentage of children with vitamin A deficiency (retinol binding protein <0.70 μmol/L, adjusted for inflammation) was 31%, 18% and 17% for the Northern, Middle, and Southern Belts, respectively. Among non-pregnant WRA (15-49 y), the prevalence of iron deficiency (serum ferritin <15 μg/L, adjusted for inflammation) followed the same pattern as that for pre-school children, with the Northern Belt (22%) being worst off, compared with the Middle (11%) and Southern (14%) Belts. Approximately half of women (50.8% in the Northern belt) had serum folate concentrations < 10 nmol/L [8].

Wheat flour fortification (iron, zinc, vitamin A, folic acid, and other B vitamins) and cooking oil fortification (vitamin A) are mandatory in Ghana; however, achieving and sustaining high compliance with target fortification levels is a challenge [9]. The 2017 Ghana Micronutrient Survey found that only 56% of oil samples (36% in the Northern Belt) were adequately fortified with vitamin A (≥10 mg/kg vitamin A), and only 6% of wheat flour samples (6% in the Northern Belt) had adequate iron concentrations (≥ 58.5 mg/kg as mandated by Ghana’s food fortification standards) [10]. Similar challenges with implementation of fortification programs have been observed elsewhere in the region [11]. Further efforts are thus required to address the high prevalence of micronutrient deficiencies in Ghana and elsewhere in West Africa.

Bouillon cubes are widely consumed in West Africa and therefore hold great potential for addressing micronutrient deficiencies in the region [12–14]. Bouillon is particularly attractive as a food vehicle because consumption is high even among subsets of the population typically at risk for micronutrient deficiencies, such as those residing in rural areas and households with low socioeconomic status [12, 15]. Estimates of the proportion of households consuming bouillon in the past week have ranged from 79% to 99% in Burkina Faso, Cameroon, Niger, and Senegal [16]. In Ghana, preliminary analyses of household data [17] suggested that 72% of households had acquired bouillon products in the prior 18 days.

In West Africa, some brands of bouillon have been produced with iodized salt since the 1990s, and bouillon products fortified with iron or vitamin A, in addition to iodine, have been introduced by some companies on a voluntary basis [18]. A study in Senegal found that iodine fortification of bouillon could potentially contribute substantially to iodine intake, by providing between 7%-115% of the recommended daily intake, depending on the iodine content of the cubes [19]. In northern Ghana, a study of schoolchildren 6-13 years of age [20] found that around two-thirds (2/3) of dietary iodine was obtained from iodine-fortified bouillon cubes. Other modelling studies have found that bouillon cube fortified with vitamin A could reduce the prevalence of inadequate vitamin A intakes without leading to an increase in excessive intakes among women or children in Cameroon [21], and would complement other intervention programs in reducing inadequate intake of vitamin A [22].

The foregoing evidence suggests that the fortification of bouillon, in combination with other ongoing measures, could contribute to the prevention of micronutrient deficiencies in Ghana. However, the levels of micronutrients that are currently included in commercial bouillon products on a voluntary basis (e.g., iron) may not be sufficient to meet public health needs [23]. Moreover, no commercial bouillon products have been released that address deficiencies in other key nutrients, such as zinc, folate, and vitamin B-12. Therefore, research is needed to explore the efficacy of fortified bouillon for addressing micronutrient deficiencies. To our knowledge, no studies, in West Africa or elsewhere, have tested the impact of multiple micronutrient-fortified bouillon on micronutrient status.

### Study objectives

The primary objectives are to assess the effects of household use of multiple micronutrient-fortified bouillon cubes (containing iodine in addition to vitamin A, folic acid, vitamin B12, iron, and zinc), compared to control bouillon cubes (fortified with iodine only), on micronutrient status and hemoglobin concentrations among non-pregnant, non-lactating women 15-49 years of age (referred to hereafter as women of reproductive age, WRA) and among children 2-5 years of age, and breast milk micronutrient concentrations among non-pregnant, lactating women 15-49 years of age at 4-18 months postpartum (hereafter, lactating women).

Secondary objectives include the effect of the multiple micronutrient-fortified bouillon, compared to the control bouillon, on the prevalence of micronutrient deficiencies and anemia; dietary intake of bouillon and micronutrients; inflammation, malaria, and morbidity symptoms; and children’s anthropometric measures and child development (details of primary and secondary outcomes for each physiological group are listed in supplemental material **S1 and S2**). Sub-studies will also assess opinions, attitudes, and acceptance (OAA) of the multiple micronutrient-fortified bouillon and hypothetical willingness-to-pay (WTP) for multiple micronutrient-fortified bouillon.

We generally hypothesize that micronutrient status, hemoglobin concentrations, dietary nutrient intake, child growth, and child development scores will be greater, and that prevalence of each micronutrient deficiency, anemia, and inflammation and morbidity will be lower, among participants who receive the multiple micronutrient-fortified bouillon compared to those who receive the control bouillon. Specific hypotheses will be reported in statistical analysis plans that will be posted prior to data analysis.

## Materials and Methods

The trial was registered on ClinicalTrials.gov (NCT05178407) and the Pan-African Clinical Trial Registry (PACTR202206868437931). The full study protocol and statistical analysis plans are posted on Open Science Framework (https://osf.io/t3zrn/).

### Study design

This community-based trial will follow a randomized, controlled doubly-masked design. WRA (n=1028), children (n=690), and lactating women (n=690) will be recruited from households in selected communities within the Kumbungu and Tolon districts, Northern Region, Ghana. The flow of study activities for each of the 3 physiological groups, and summary of exclusion and deferral criteria, are illustrated in **Figure 1**, while the schedule of data collection is summarized in **Table 1**. Written informed consent will be obtained from participating women and from caregivers of participating children. Consented WRA will be invited for a screening visit for assessment of total body vitamin A stores using the retinol isotope dilution (RID) technique [24]. Following a 3-week observation period, which includes 2 in-home visits, all participants will undergo baseline assessment, and eligible participants will be enrolled by being randomly assigned to receive household rations of one of two types of bouillon cubes: 1) a multiple micronutrient-fortified bouillon cube containing vitamin A, folic acid, vitamin B12, iron, zinc, and iodine, or 2) a control cube fortified only with iodine. Households will be advised to prepare their meals as usual, but will be requested to use the study-provided bouillon instead of the commercially available bouillon they typically purchase. Households will then be visited every 2 weeks for delivery of study cubes, monitoring of adherence, and recording of morbidity symptoms. Endline assessments will be conducted after ∼9 months (35 and 38 weeks) for WRA and children, and after ∼3 months (11 and 12 weeks) for lactating women. Details of study procedures and data collection at each time point are summarized in **Table 1** and below.

**Figure 1.** Flow of study activities.

**table 1.**
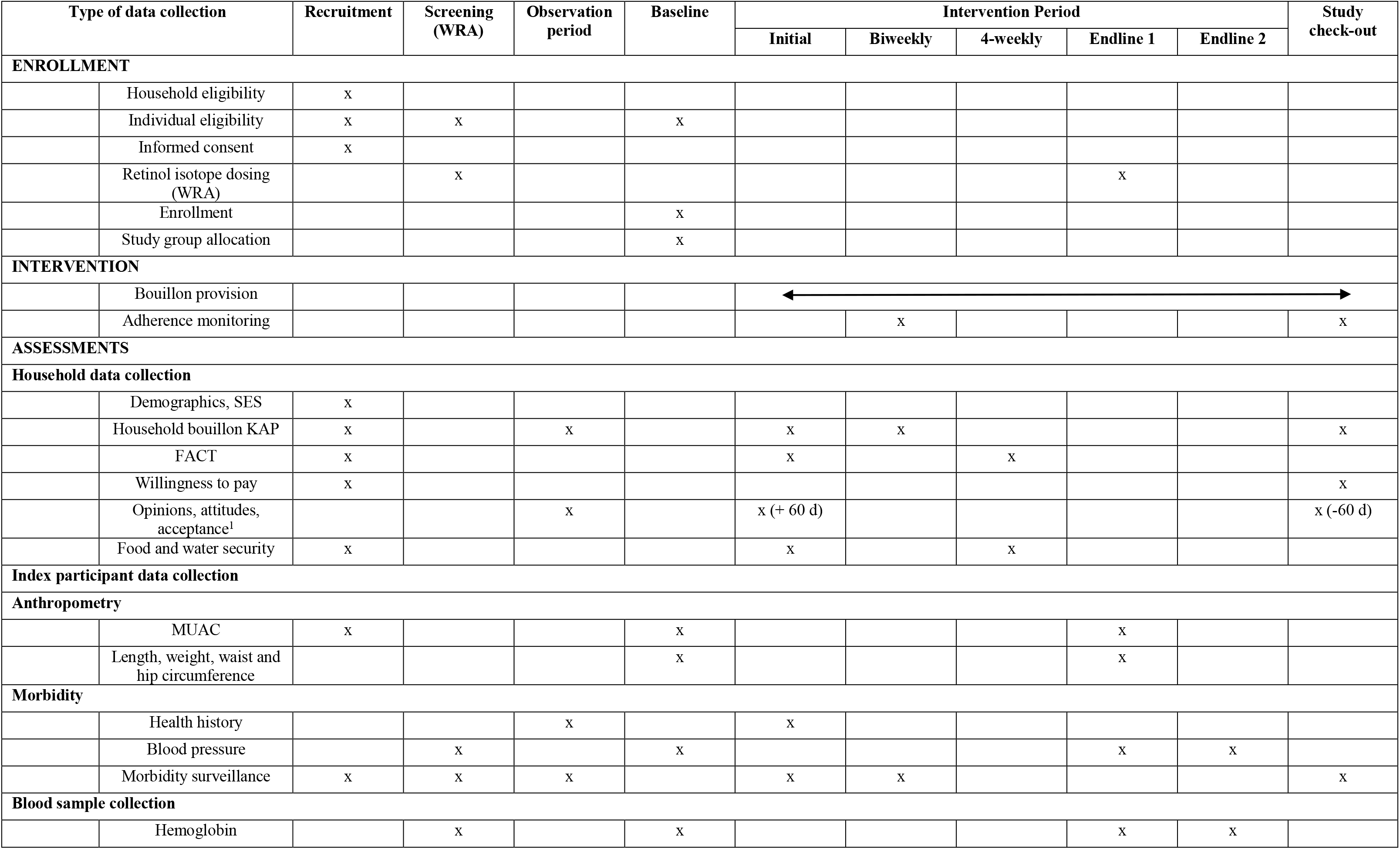
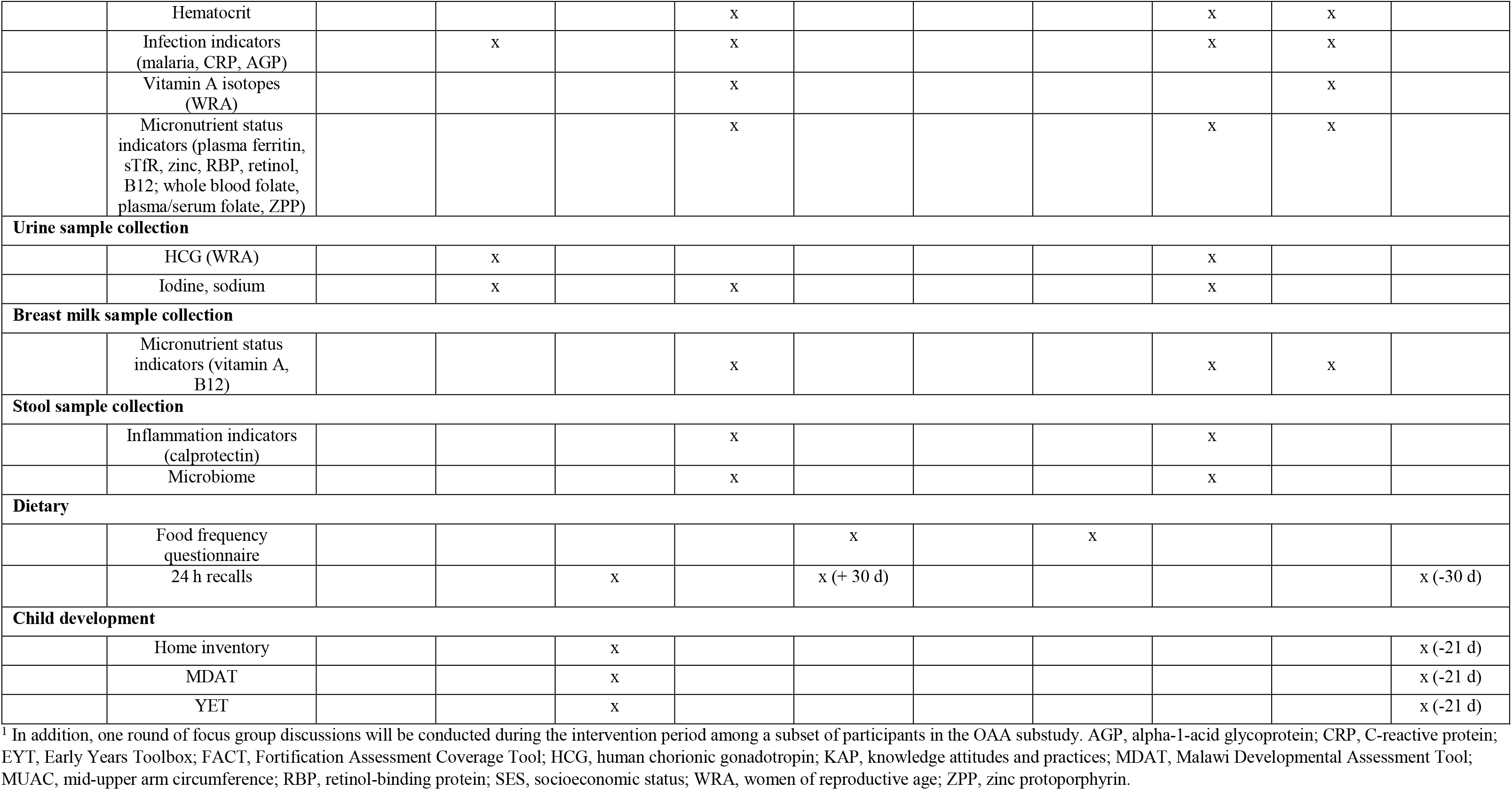
Schedule of recruitment, screening, enrolment, intervention, and assessments.

### Study site

This proposed research will take place in the Tolon and Kumbungu districts in the Northern Region. Micronutrient biomarker data from a pilot survey in these districts (not yet published) confirmed prior research [10] that suggests micronutrient deficiencies are common. For example, pilot data indicated that 28% of WRA and 53% of children 2-5 years of age were iron deficient (serum ferritin concentration <15 or <12 µg/L, respectively, adjusted for inflammation). In addition, 34% of children had evidence of vitamin A deficiency (serum retinol <0.70 µmol/L, adjusted for inflammation) although only ∼1% of WRA had serum retinol <0.70 µmol/L. Serum vitamin B12 concentrations were low (<221 pmol/L) among 12% of women and 19% of children 2-5 y, while low vitamin B12 concentrations in breast milk (<221 pmol/L) were observed among 40% of lactating women. Anemia was also common among both women (31%) and children 2-5 years of age (36%). Consumption of micronutrient supplements of any type was rare, with fewer than 10% of WRA and children interviewed in the pilot survey reporting consumption in the previous 30 days. Similarly, pilot survey data also confirmed inadequate compliance with mandatory target micronutrient fortification levels of industrially fortified foods. Data on micronutrient levels in food samples collected at the study site indicate that oil samples were fortified with amounts equivalent to ∼69% of target vitamin A levels and wheat flour samples were fortified with ∼33-34% of target levels for iron and zinc (not yet published).

### Identification and recruitment of participants

Participants in the study will be 1) non-pregnant non-lactating WRA 15-49 years of age, 2) children 2-5 years of age, and 3) non-pregnant, lactating women 15-49 years of age at 4-18 months postpartum. Multiple participants may be recruited from the same household if they represent different physiological groups (for example, a WRA and child 2-5 years of age), but only one index participant (e.g., one WRA) from each physiological group will be recruited from each household. A household is defined as a group of people who recognize the same head of household, live together, and share living expenses and meals (see **S5. Supplemental Methods** for full definition). Sub-study respondents will be 1) OAA: an adult household member (≥ 15 years of age) who is not recruited in the trial, and 2) WTP: the household member who is primarily responsible for making decisions about bouillon purchasing.

Within the Tolon and Kumbungu districts, we will recruit potential participants from 16 recruitment sites composed of 14 “rural” (comprising both rural and semi-urban areas) and 14 urban Ghana Statistical Service designated sampling areas. Recruitment of each physiological group (WRA, children, lactating women) will be balanced within each recruitment site so that the final sample of participants in each physiological group represents a similar geographic area.

Households in selected recruitment sites will be identified through a random walk method with door-to-door recruitment[25]. Trained enumerators fluent in English and the local language (Dagbani) will approach households to assess eligibility, administer recruitment questionnaires, and conduct the informed consent process.

### Consent

Upon selecting a household to approach, enumerators will first obtain oral consent from the head of household to proceed with the interview and then will administer a series of questionnaires to assess interest in study participation and complete a household roster. The household roster will be used to identify potentially eligible individuals. If more than one potentially eligible individual per physiological group is present in the home (for example, if there are two children between 2-5 years of age), one individual per physiological group will be randomly selected based on a Kish Table [26]. Potentially eligible participants will then complete the written informed consent process, wherein enumerators read aloud the consent form in the preferred language of the participant (English or Dagbani), concluding with an informed consent quiz to confirm the participant’s understanding of the study procedures. Informed consent will be confirmed with a signature or thumbprint; impartial witnesses (community members not affiliated with the study) fluent in English and Dagbani will be present for the consent process for non-literate participants to ensure the information presented orally matches the written informed consent sheet. Caregivers will provide consent for children 2-5 years; participants 15- 17 years who have never married and are living with their parents will provide assent and their caregiver will provide consent. After an index participant has consented to participate in the study, respondents for the OAA and WTP sub-studies will be recruited. For the OAA sub-study, enumerators will use the household’s roster and a random number generator to randomly select the OAA participant from all remaining adult household members (e.g., ≥15 years and not recruited into the RCT); one OAA participant will be recruited per household. Upon agreement to participate in the OAA sub-study, the selected adult household member will provide written informed consent. For WTP, the household member with primary responsibility for bouillon purchasing (who may or may not be a participant in the RCT) will be asked to provide oral consent to respond to questions to elicit their hypothetical willingness to pay for a fortified bouillon cube.

### Inclusion and Exclusion Criteria, and Deferral Criteria

Detailed descriptions of the research inclusion and exclusion criteria by physiological group and at each stage of the study are included in **S5. Supplemental Methods** and summarized in **Figure 1**. Briefly, households will be eligible for recruitment if the household is willing to use the study-provided bouillon cubes for the duration of the study and no one in the household i) is currently participating in another research trial, ii) has a chronic medical condition requiring frequent blood transfusions, or iii) has an allergy to ingredients in the bouillon cubes (shrimp, wheat, milk, soy, eggs, celery, fish, or mollusc). WRA, lactating women, and children will be eligible for inclusion if they i) do not have a severe acute or chronic illness, ii) are able to provide informed consent, and iii) plan to remain in the study area for the duration of the study.

WRA must have a negative malaria rapid diagnostic test (RDT) result and C-reactive protein (CRP) <5 mg/L at the RID screening visit, and hemoglobin (Hb) ≥8 g/dL at the screening and baseline visits. Children must have mid-upper arm circumference (MUAC) of ≥11.5 cm at recruitment, and Hb ≥7 g/dL, weight-for-height z-score (WHZ) ≥-3 SD, and no bilateral pitting oedema at baseline.

Throughout the study, WRA and lactating women who report pregnancy, and lactating women who report lactation cessation, will be excluded. Participants who present with COVID-19- related symptoms, or malaria or elevated CRP (WRA screening visit only), will be required to defer study assessments for 2-3 weeks (**Figure 1; Supplemental Methods**). At the RID screening or baseline visits, if participants do not meet inclusion criteria after the deferral period, they will be excluded.

The following sections describe the sequence of study visits and procedures.

### Screening and baseline visits

Following recruitment and consent, WRA will visit the study site to be screened for eligibility for the RID test for assessment of total body stores of vitamin A [24]. Then, during a 3-wk observation period, descriptive information will be collected for all participants at two in-home visits. Following the 3-wk observation period, all participants will visit the study site for baseline assessments; eligible participants will be randomized to a treatment group. Procedures at each visit are summarized below.

#### RID screening visit for non-pregnant, non-lactating women of reproductive age

WRA will be screened for eligibility for the RID test using procedures described below. Fieldworkers will assess participants for recent morbidity, vitamin A intake (e.g., liver or supplements in the past 24h) and pregnancy (based on human chorionic gonadotropin in urine), measure blood pressure and collect a capillary blood sample. WRA with no recent (past 72h) morbidity or intake of vitamin A-rich foods (e.g., liver) in the past 24h, who have a negative pregnancy test, Hb ≥8 g/dL, CRP <5 mg/L, and a negative malaria RDT result will receive an oral dose of d6-labelled vitamin A (2 mg retinol equivalent, sourced from Buchem BV, Apeldorn, Netherlands) in sunflower oil. The labelled vitamin A will be certified as food grade (Food Chemical Codex IV) and approved by the Ghana Food and Drugs Authority. All WRA who received the d6-labelled vitamin A dose will be invited to the biospecimen collection site 21 days after the isotope dose (based on Green et al., 2021 [27]) for a venous blood sample to assess total body vitamin A stores using the RID technique. This 21-day visit will be scheduled to coincide with the trial baseline visit (procedures described below).

#### Observation period

For all participants, during the ∼3-week period between recruitment and the baseline visit (i.e., the observation period), information on participant and household characteristics, dietary intake, morbidity, and bouillon use will be collected during two household visits conducted 2 weeks apart. Baseline child development assessments will also occur during this period. Additionally, OAA participants will complete a demographic questionnaire and one quantitative survey about opinions, attitude, and acceptance of commercial bouillon.

#### Baseline visit

After completion of the observation period, eligible participants will be invited to the mobile biospecimen collection site for the baseline visit (**Figure 1**; **Table 1**). Following morbidity screening, participants will undergo baseline data collection procedures, including anthropometry, measurement of blood pressure and collection of a spot urine sample (WRA and children only), and collection of a stool sample (children only). Venous blood will be collected from WRA (fasted) and children (nonfasted) for assessment of hemoglobin, malaria, and biomarkers of micronutrient status and inflammation. For lactating women, a full breast milk sample will be collected for micronutrient analysis. Questionnaires will be used to collect individual level information related to the interpretation of biospecimen results.

Following the baseline data collection procedures, households with participants who remain eligible for the study will be randomized to a study arm; an enumerator will visit the household on the following day (the initial home visit) to deliver the first ration of bouillon cubes that corresponds to the assigned treatment group.

### Randomization and blinding

The unit of randomization will be at the household level, regardless of the number of individual participants (up to 3) eligible within that household. To randomize the households, the trial statistician will create a computer-generated block randomization scheme with randomly selected block lengths of four or eight. Pieces of paper bearing intervention group assignments represented by 4 different 3-digit alphanumeric codes (2 codes per study arm) and numbered 1– 2408 (maximum number of households if each participant resides in a different household) will be placed in opaque envelopes and stacked in increasing order by study staff not involved in the randomization process. For each household enrollment, a designated and trained study personnel will select the four topmost envelopes from the stack and ask the participant to choose an envelope to reveal their study group assignment. At this point, the household ID (and the participant IDs for each eligible participant within the randomized household) will be linked to one of the four study codes indicating the intervention the household will receive.

All study bouillon cubes will be individually wrapped in identical packaging and will be identified only by the study code (a 3-digit alphanumeric code). Boxes of cubes will be labelled with the study code and an associated group color. Participants and study personnel will know the code (and for study personnel, the group color) to which each household is assigned but will not know the micronutrient content of the products.

Two code keepers (one at the University of California, Davis, UCD, and one at the University of Ghana) who are not otherwise affiliated with the study will be responsible for assigning the 4 study codes to the intervention products and communicating directly with the manufacturer about the group assignments. In addition, a hard copy of the randomization code will be stored in a sealed envelope in a locked filing cabinet at the University of Ghana. Two individuals, unaffiliated with the study, will have a key to the office and access to the key to the filing cabinet. In the case of a severe adverse reaction pertaining to an individual participant, the blinding code will be broken only to identify the bouillon cube formulation that the individual participant received, if necessary for their medical care. Only individuals directly involved in the care of the study participant will be unblinded on a strict need-to-know basis.

The specific bouillon cube formulation associated with each of the 4 study codes will also be stored in sealed envelopes held by the co-principal investigators and the statistician. The envelopes will be opened only after statistical analyses of the primary outcomes are completed and the investigators have reached consensus on interpretation of the results.

### Intervention period procedures

Enumerators will visit participant households every 2 weeks throughout the study to provide the study bouillon rations and collect information on adherence (e.g., household inventory of study bouillon and commercial bouillon, bouillon wrapper counts, bouillon use) and morbidity. All reasonable efforts will be made to complete follow-up assessments, regardless of adherence.

Questionnaires to collect information on household food insecurity, water insecurity, consumption of fortified foods, and dietary patterns (e.g., dietary diversity and child feeding) will also be administered during the initial home visit and every 4 weeks thereafter (Table 1). Cube delivery and data collection will be conducted with “no contact” methods (e.g., phone interview) in the case of COVID-19 exposure or symptoms. Baseline dietary assessment will be conducted by administering two 24-hour dietary recall interviews per participant on non-consecutive days within 30 days after the baseline visit (all participants), and two additional 24- hour dietary recalls will be conducted on non-consecutive days within the 30 days before the second endline visit (WRA and children only).

Within the first two months of the intervention period, OAA participants will complete a similar quantitative survey to that administered in the observation period but focused on the study-supplied bouillon cubes, and again within the last two months of the intervention period. OAA participants in households in which the only trial participant is a lactating woman will complete only one intervention period survey.

### Endline visits

At the end of the intervention period, participants will be invited to the mobile biospecimen collection site for the two endline data collection visits (35 weeks and 38 weeks for WRA and children; 11 weeks and 12 weeks for lactating women). A 3-week period between endline visits was chosen based on the desired timing between the isotope dose administration and blood collection for assessment of total body vitamin A stores. Delivery of study bouillon cubes will continue through the last endline assessment for each household. Data collection procedures at the first endline visit will be similar to those described at the baseline visit. Briefly, data collection will include anthropometric measurements and morbidity symptoms (all participants); spot urine samples, blood pressure measurement and venous blood collection (WRA and children); breast milk collection (lactating women); and stool collection (children).

In addition, at the first endline visit (35 wk), WRA will receive an oral dose of ^13^C_10_-retinyl acetate (2 mg retinol equivalents, sourced from Buchem BV, Apeldorn, Netherlands) to assess total body vitamin A stores. ^13^C_10_-retinyl acetate will be used at endline to avoid any potential interference from the baseline dose of d6-retinyl acetate. To assess eligibility for the endline isotope dose, participants will undergo screening procedures identical to those at the pre-intervention screening visit, including assessment of morbidity symptoms, vitamin A intake, pregnancy (based on measurement of HCG in urine), anemia, malaria infection, and inflammation. Based on results of these assessments, WRA will either be 1) eligible to receive the endline isotope dose and continue with the study; 2), deferred and scheduled to be re-screened for isotope dosing at the 2^nd^ endline visit (recent morbidity, CRP > 5 g/L, positive malaria RDT); 3) excluded from isotope dosing and the second endline blood collection (participants with Hb < 8 g/dL), or 4) excluded from the study (positive pregnancy test) (see **S5. Supplemental Methods** for detailed exclusion and deferral criteria).

Data collection procedures at the second endline assessment will include a subset of the outcomes measured at the first endline assessment, measured using identical methods. These will include morbidity symptoms (all groups), venous blood collection (WRA and children) and breast milk collection (lactating women). WRA who are deferred at the first endline visit, and do not receive the vitamin A isotope dose, will be re-screened at the second endline visit using the same procedures for eligibility described above. WRA who are eligible to receive the isotope dose at the second endline will be asked to return for an additional endline visit to collect a follow-up venous blood sample for RID assessment 3 weeks later.

In between the two endline visits (35wk and 38wk), a single endline child development assessment will be completed for each task. After the second endline visit, a final home visit will be conducted to complete the study exit form and collect all remaining study bouillon cubes from the household. In each household, the baseline assessment of hypothetical willingness-to-pay for a fortified bouillon cube will be repeated at endline.

### Intervention: Fortified bouillon development

#### Product formulation

Bouillon cubes for the trial will be produced based on a non-proprietary formulation that is comparable to commercial cubes sold in Ghana, containing ∼52% salt by weight, along with palm fat, corn starch, spices and flavour enhancers, colouring, and micronutrient premix (Nicholas Archer, CSIRO, personal communication). Shrimp flavour was selected, as it was the primary bouillon flavour used by all households in the pilot survey in the study community [28]. **Table 2** presents the micronutrient concentrations, chemical fortificant forms of each nutrient, and equivalent daily target micronutrient doses for women and young children in the active arm of the study. The specific fortificants were selected on the basis of technical compatibility with the food matrix and experience with use as fortificants [29]. In the case of iron, the combination of ferric pyrophosphate (FePP) with a citric acid/trisodium citrate buffer (CA/TSC) is expected to enhance bioavailability compared to FePP alone [30].

**Table 2.** Target fortification levels, estimated daily dose of micronutrients delivered by fortified bouillon, and percent of the recommended daily allowance for women of reproductive age and young children^1^.

| **Micronutrient (Fortificant)** | **Amount of micronutrient per gram bouillon<sup>2</sup>** | **Daily amount provided to women<sup>3</sup>** | **Daily amount provided to women, as proportion of the RDA** | **Daily amount provided to children** | **Daily amount provided to children as proportion of the RDA** |
| --- | --- | --- | --- | --- | --- |
| Vitamin A (retinyl palmitate) | 200 µg | 400 - 500 µg | 57 - 71% | 200 µg | 50 - 67% |
| Folic acid | 80 µg | 160 - 200 µg | 67 - 83% | 80 µg | 67 - 89% |
| Vitamin B12 | 1.2 µg | 2.4 - 3.0 µg | 100 - 125% | 1.2 µg | 100 - 133% |
| Iron (FePP/CA/TSC) | 4 mg | 8 - 10 mg | 44 - 56% | 4 mg | 40 - 57% |
| Zinc (ZnO) | 3 mg | 6 - 7.5 mg | 75 - 94% | 3 mg | 60 - 100% |
| Iodine (KIO<sub>3</sub>) | 30 µg | 60 - 75 µg | 40 - 50% | 30 µg | 33% |

<sup>1</sup>Recommended Daily Allowance (RDA) values are for women 19-50 years of age and children 1-3 y and 4-8 y [33]. FePP/CA/TSC: ferric pyrophosphate/citric acid/trisodium citrate. Values do not include overage. Overages based on expert opinion were added to account for losses during storage and cooking (30% vitamin A, folic acid, and vitamin B12; 20% iodine; 0% iron and zinc).
<sup>2</sup>Values represent the amount of the active form of the nutrient, e.g. 200 µg retinol equivalents as retinyl palmitate and 4 mg iron as FePP/CA/TSC.
<sup>3</sup>Range of values reflects assumed bouillon cube intake of 2-2.5 grams per day among WRA. For children, calculations assume 1 gram per day and RDA categories of 1-3 years and 4-8 years of age (representing children 2-5 years of age).

With estimated typical bouillon consumption of 2-2.5 g/d among WRA and 1 g/d among children, these cubes supply a daily dose of micronutrients equivalent to 33% to 133% of the corresponding US Recommended Daily Allowance for the 6 micronutrients added to the cubes (**Table 2**). The concentrations of micronutrients were selected based on: 1) modelling of national survey data from Ghana and other West African countries to estimate the contribution of hypothetical fortification of bouillon to adequacy of dietary micronutrient intake (to estimate the potential public health benefit) [14]; 2) the estimated amount of bouillon consumed in the communities selected for the study; 3) expected efficacy, based on a literature review of the effect of micronutrient interventions on change in micronutrient status as measured by biomarkers or on clinical outcomes (anemia and neural tube defects) and estimated accumulation of micronutrient stores (vitamin A and iron); and 4) safety considerations, including the likelihood of study participants or their household members exceeding the tolerable upper intake level (UL), and a review of the literature to identify adverse effects of micronutrient interventions at varying daily micronutrient doses. Additional details on the rationale for the selected fortification levels and estimated daily micronutrient doses are provided as a Technical Appendix to the study protocol (available at https://osf.io/t3zrn/).

The control cube formulation contains iodine but no other added micronutrients. This selection of micronutrient for the control cube is consistent with current policy in Ghana, where salt iodization is mandated (15 mg/kg at household level) [31] and commercial bouillon cubes are typically made with iodized salt [18].

#### Product testing

Acceptability of the unfortified, non-proprietary cube formulation was confirmed through consumer testing in northern Ghana, including using commercial cubes as a reference (Nicholas Archer, CSIRO, personal communication). The effects of micronutrient fortification on micronutrient stability and sensory properties of the multiple micronutrient-fortified bouillon were evaluated in accelerated storage trials. The results confirmed good stability and cube quality after 2 weeks of storage of bouillon cubes (in individual wrappers) under accelerated storage conditions (37°C, 75% relative humidity), and for at least 5 months for storage under optimal conditions (23°C, 50% relative humidity) (Nicholas Archer, CSIRO, personal communication). Finally, acceptability of the multiply-fortified and control cube formulations used in this study was evaluated among a sample of women and their households in the study community [32]; overall, the study results suggested that the bouillon formulations were well liked and would be suitable for use in further research studies.

#### Bouillon cube production and management

Bouillon cubes will be produced at industrial scale by GB Foods (Barcelona, Spain). Bouillon cubes will be shipped by air or sea to Ghana and transported to a dedicated storage facility at the study site. Boxes of bouillon will be stored on pallets in a cool (12 – 16° C) temperature-controlled environment with frequent temperature and humidity monitoring. Micronutrient content of the micronutrient premix and the study bouillon cubes will be analyzed to ensure proper coding and formulation of the bouillon cubes before distribution to participants.

#### Household bouillon ration

The household bouillon ration will be determined based on household size and estimated bouillon consumption (2.5 g/capita/d). This approach was based on previous research in the study community showing that median per household bouillon consumption was 20 (12, 24) g/d (median, p25, p75) and that bouillon consumption was 2.4 g/d among WRA and 1.0 g/d among children, similar to the assumptions in Table 2 [28]. The household bouillon ration may be revised over the course of the study based on observations and questionnaire responses (e.g., changes to household composition) but will not exceed 4 g/capita/d. Bouillon rations will be delivered by enumerators to participants’ homes every 2 weeks and stored in study-provided plastic containers. Households will be encouraged to use the study bouillon (in lieu of commercially purchased bouillon cubes) according to their typical cooking habits. At the initial home visit (first day of bouillon distribution) only, the enumerator will request to collect all commercial bouillon in the household; if the household agrees, they will be compensated according to a table of current brand-specific market prices for bouillon. During the trial, households are free to purchase and use commercial bouillon cubes as they choose.

### Data collection methods

#### Questionnaires

Trained enumerators will administer all questionnaires in the preferred language of the participant (English or Dagbani) as structured oral interviews using electronic tablets. Data on demographic and socio-economic status (household size and composition, assets, housing quality, sanitation and hygiene, and individuals’ education and occupations), food security [Household Food Insecurity Access Scale (HFIAS) [34]] and water security [Household Water Insecurity Experiences (HWISE) [35]] will be collected. Pilot-tested questionnaires will also collect information on household bouillon acquisition, consumption, and stocks; bouillon knowledge, attitudes and practices (KAP); consumption of fortified foods, including bouillon [Fortification Assessment Coverage Tool (FACT)[26]; and hypothetical willingness-to-pay (WTP) for a new micronutrient-fortified bouillon cube. Information on health history, non-communicable disease risk [WHO STEPwise approach to NCD risk factors surveillance (STEPS) questionnaire] [36], and breastfeeding and infant and young child feeding practices (IYCF) [37] and dietary diversity (Dietary Quality Questionnaire [DQQ]) [38] will also be collected. Parent perception of child weight and parental beliefs, attitudes and practices toward child feeding will be assessed with a child feeding questionnaire (CFQ) [39]. Structured questionnaires will be used to assess recent morbidity (presence and severity of selected symptoms of pre-defined diseases and serious adverse events).

#### Adherence

Households will be requested to save wrappers from all bouillon (both study-provided and commercial) used in the household since the last biweekly visit. To monitor adherence, at each biweekly visit during the intervention period, enumerators will count the bouillon wrappers and record an inventory of all bouillon cubes in the household (both study-provided and commercial). In addition, enumerators will administer a questionnaire to gather data about household purchases of bouillon and household bouillon use in the past 2 weeks, including sharing of study bouillon cubes with non-household members. Households with sustained low adherence (i.e., low consumption of study-provided bouillon) will be identified through routine adherence data monitoring; enumerators will be instructed to inquire about the reasons for low adherence and to encourage use of the study bouillon.

#### 24 hour recalls

Quantitative dietary intake data will be collected through 24-hour dietary recall interviews to capture bouillon and nutrient intake both prior to and while receiving the study cubes. All participants (WRA, lactating women, children) will complete one dietary recall during the observation period and two baseline dietary recalls on non-consecutive days within the first 30 days of the intervention period. WRA and children will complete two additional endline recalls on non-consecutive days within the 30 days before the second endline visit. Lactating women will not complete endline dietary recalls due to the shorter study duration.

Dietary recalls will be conducted by enumerators with extensive training and practice on the 24- hour recall technique. Interviews will follow a multiple pass methodology [40] to collect information on all foods and beverages consumed by each participant during a defined 24 hour period. Recall data will be collected using a tablet-based data collection system (an adaptation of OpenDRS [41]) that allows for direct entry of dietary data and recipe collection on the tablet, through a platform that guides interviewers through the dietary recall interview and suggests prompts to aid in the interview process. Dietary recalls will be conducted in participant households to take advantage of the opportunity to use the participants’ typical utensils for portion size estimation where possible. Enumerators will also carry “kits” with common utensils and foods (e.g., salt, sugar, spices, etc.) to obtain weights for portions and ingredients where possible.

#### Opinions, attitude, and acceptance (OAA) towards the study bouillon among adult household members

The OAA sub-study will employ a mixed methods approach (quantitative survey and focus group discussions). OAA towards commercial bouillon will be evaluated with a questionnaire during the observation period, and towards the study-provided bouillon with a similar questionnaire once within the first 60 days and once within the final 60 days of the intervention period. One round of focus group discussions (FGDs) will also be conducted among a subset of participants who complete the OAA questionnaires to qualitatively assess OAA of the study-supplied bouillon cubes. FGDs will be conducted in Dagbani following a pre-tested FGD guide and audio-recorded.

#### Anthropometry

Anthropometric data will be collected by trained, standardized anthropometrists, following protocols established by the Food and Nutrition Technical Assistance Project [42]. Children’s MUAC (left arm; UNICEF S0145620), standing height of all participants (SECA 217), and the waist and hip circumference of WRA (SECA 201) will be measured to 0.1 cm precision. Weight of all participants (lightly clothed) will be measured to 50 g precision (SECA 874). All measurements will be collected in duplicate, and the average of the two measurements calculated per participant for each outcome. If the two measurements differ by more than 0.1 kg (weight), or 0.5 cm (MUAC, height, waist or hip circumference), the measurement will be repeated, and the two closest measurements will be retained for data analysis.

#### Child development

Child development will be assessed with the Malawi Development Assessment Tool (MDAT) [43], which measures four domains of development (gross motor, fine motor, language, and personal-social), and the tablet-based Early Years Toolbox Go-No-Go Task which measures executive function (EYT) [44]. Assessments will take place at a central data collection site or (if necessary) in a low-distraction location at the participant’s home. Characteristics of the home environment and caregiver-child interaction will be assessed at a household visit with the Home Observation Measurement of the Environment (HOME) tool [45]. To assess the test-retest reliability of the EYT a convenience sample of 30 children will be asked to come back 1 week later to repeat the EYT.

Prior to data collection, enumerators trained in child development assessment will be required to take a written and practical assessment on the data collection procedures. Inter-rater reliability of the enumerators will be assessed for the MDAT and the HOME to ensure consistency of data collection. The MDAT and HOME were previously piloted in the study site and found to be feasible for use in this setting.

#### Biological sample collection and laboratory analyses

##### Capillary and venous blood collection

At the pre-intervention RID-screening visit, capillary blood samples will be obtained from WRA to measure hemoglobin and CRP concentrations using the QuikRead Go (Aidian Oy, Espoo, Finland); malaria antigenemia will be measured by RDT (BioZEK, B.V., Apeldoorn, Netherlands). At the post-intervention RID-screening visit, these measurements will be done using venous blood that is collected for assessment of endline micronutrient status.

At baseline and endline assessments, venous blood samples will be collected from children and WRA by trained laboratory technicians, following standard biological safety protocols and procedures recommended by the International Zinc Nutrition Consultative Group (IZiNCG) [46]. Samples will be collected at a central location in each community (mobile biospecimen collection site) in the early morning. WRA, but not children, will be asked to fast for at least 8 hours prior to their blood draw appointment; time of blood collection and last meal will be recorded for all participants. At all blood sample events (baseline, endline 1 and endline 2), 7.5 ml of blood will be drawn from the antecubital vein into an evacuated, trace element-free polyethylene blood collection tube containing lithium heparin (Sarstedt AG & Co, Numbrecht, Germany; ref 01.1604.400). For all WRA at baseline and endline 2, as well as a subset of WRA at endline 1, an additional 4.5 ml of blood will be drawn from the antecubital vein into an evacuated serum blood collection tube (Sarstedt AG & Co, Numbrecht, Germany; ref 05.1104.100); this will occur prior to blood collection in the lithium heparin tube. In addition, if WRA receive the vitamin A isotope dose at endline 2 (due to deferral), an additional 4.5 ml of blood will be drawn from the antecubital vein into an evacuated serum blood collection tube (Sarstedt AG & Co, Numbrecht, Germany; ref 05.1104.100) at a third time point. If the phlebotomist is unable to obtain a venous blood sample after two attempts, a capillary sample will be obtained by fingerstick and collected into a 200 µl microvette containing lithium heparin (Sarstedt AG & Co, Numbrecht, Germany; ref 20.1292.100). Following blood collection, all blood collection tubes will be placed into a battery-operated cold box (4 – 8°C) until processing.

Immediately following blood collection, point-of-care assessments will be performed using whole blood remaining in the butterfly tubing or needle (or from the capillary sample); if necessary, whole blood may be aliquoted from the lithium heparin blood collection tubes for the RDTs. Hemoglobin will be measured using a portable photometer (Hemocue 301, HemoCue AB, Angelholm, Sweden). Malaria antigenmia will be assessed with an RDT. Hematocrit (WRA only) will be measured using the GCH-24 Hematocrit centrifuge (Globe Scientific, Mahwah, New Jersey, USA). In addition, hemoglobin, hematocrit and zinc protoporphyrin (ZPP) will be measured using the iCheck Anemia point-of-care device (Bioanalyt, Teltow, Germany).

Whole blood samples from the serum blood collection tubes will be allowed to clot for 30-60 minutes, after which time they will be centrifuged at 1097 x g (3100 RPM) for 10 minutes (PowerSpin Centrifuge Model LX C856E; United Products & Instruments, Inc, Dayton, NJ). Serum will be aliquoted for the measurement of concentrations of [^12^C]retinol, d6-retinol and [^13^C10]retinol and folate. From the lithium heparin blood collection tube, 100 µl of whole blood will be aliquoted into a cryovial containing 1 ml of 1% ascorbic acid solution for the measurement of folate concentration (to calculate erythrocyte folate) among WRA. The remaining heparinized blood will be centrifuged within 8 h of collection at 1097 x g (3100 RPM) for 10 minutes (PowerSpin Centrifuge Model LX C856E; United Products & Instruments, Inc, Dayton, NJ). Plasma will be aliquoted into cryovials for the measurement of micronutrient status (i.e., ferritin, soluble transferrin receptor [sTfR], retinol binding protein [RBP], retinol, zinc, folate and B12) and indicators of systemic inflammation (i.e., CRP, and alpha-1-acid glycoprotein, AGP).

#### Breast milk collection and processing

At baseline and endline assessments, breast milk samples will be collected from lactating women using the “full milk sample” methodology [47] using the Medela Swing single electric breast pump (Barr, Switzerland). Breast milk will be collected from the breast from which the infant has not fed for a longer time (i.e., the breast that is ‘more full’), with a minimum of one hour elapsed between the milk sample collection and previous feeding. Time of day and time since last feed will be recorded. Fat will be measured in fresh milk in triplicate using the creamatocrit method (Creamatocrit Plus Centrifuge; EKF Diagnostics) [48]. The milk sample will be mixed well and ∼6 ml will be aliquoted for the measurement of vitamins A and B12; the remainder of the milk sample will be returned to the lactating woman to feed to her child.

#### Urine collection and processing

Urine samples will be collected from both WRA and children at baseline and endline 1 assessments. Among WRA only, spot urine samples will be collected on the day of the pre- and post-intervention RID-screening visits (prior to retinol isotope dosing) to assess pregnancy status based on rapid tests for human chorionic gonadotropic (HCG). At all time points, urine will be aliquoted into cryovials for the measurement of iodine, sodium, potassium and creatinine concentrations.

#### Stool collection and processing

Stool samples will be collected from children at baseline and endline 1. The child’s caregiver will be provided with stool sample collection kits (self-standing transport tube with screw cap and spoon, absorbent pad and/or diaper, gloves, biohazard bags, soap) and requested to collect the first available fresh stool sample from the child on the morning of the baseline visit, partially filling the transport tube. In a randomly selected subset of participants, caregivers will also be provided with a stool nucleic acid collection and preservation tube (Norgen Biotek; Ontario, Cananda), and requested to collect a small sample of stool into a second tube. Unpreserved stool samples will be aliquoted into cryovials for the measurement of calprotectin; no processing will be necessary for the microbiome analysis samples in the nucleic acid collection and preservation tubes.

#### Biological sample storage

All biological sample aliquots will be stored in a battery-operated cold box (4 – 8°C) and transported daily from the biospecimen collection site to the District Hospital at Tolon for storage in a -86°C freezer, with regular temperature monitoring. Samples thus stored will be periodically transported to the University of Ghana for longer-term frozen storage before analysis. Samples to be analysed outside Ghana will be shipped by air on dry ice to the respective labs, as described below.

#### Laboratory analysis

Plasma ferritin, sTfR, RBP, CRP, and AGP concentrations will be measured in duplicate using a combined sandwich enzyme-linked immunosorbent assay (ELISA) technique at VitMin Lab (Willstaett, Germany) [49]. Serum concentrations of d6-retinol, [^12^C]retinol and [^13^C10]retinol will be measured by liquid chromatography with tandem mass spectrometry (LC/MS/MS) at Newcastle University (Newcastle, UK) [50]. Plasma [51] and breast milk [52] vitamin A concentrations will be measured by high performance liquid chromatography (HPLC) at the UCD (Davis, CA). Plasma zinc concentrations will be analyzed by inductively-coupled plasma optical emission spectrophotometry (ICP-OES; Agilent 5100 SVDV, Santa Clara, CA) at the University of California, San Francisco (UCSF) Benioff Children’s Hospital (Oakland, CA) [53]. Whole blood folate and serum/plasma folate will be analyzed by the US Centers for Disease Control and Prevention (Atlanta, GA) and used to calculate erythrocyte folate concentrations [54]. Vitamin B12 concentrations in plasma and breast milk, will be measured by the Western Human Nutrition Research Center (Davis, CA). Urinary iodine concentration will be assessed using a modification of the Sandell-Kolthoff reaction at the University of Ghana [31]. Urinary sodium, potassium and creatinine concentrations will be analyzed at the UCSF Benioff Children’s Hospital (Oakland, CA). Stool calprotectin will be analysed using a commercially available kit and gut microbiota will be characterized by bacterial 16S rRNA sequencing at UCD (Davis, CA). Novel indicators and other indicators of micronutrient status, growth and infection may be assessed if funding becomes available, among participants who consented to the use of their specimen for future research.

### Data management and analysis

#### Data management and confidentiality

Data will be collected directly on tablets via forms developed to include pre-determined logic checks and range validations using the SurveyCTO® program. Selected data collected on paper forms will be double-entered. Data will be monitored daily by field supervisors. Weekly reports for monitoring of key variables, including study enrollment, attrition, missing data/forms, adherence, and selected quality control monitoring will be routinely reviewed by the research team and field supervisors to identify action items (e.g., form corrections, refresher trainings, participant visits to encourage adherence, data corrections, etc.). Study procedures are designed so that all Personally Identifying Information is kept separately from all other data throughout data collection, storage, and analysis and only accessible to a limited number of authorized members of the research team.

#### Outcome definitions

For WRA and children, primary outcomes are hemoglobin concentrations and biomarkers of status of iron (plasma ferritin and sTfR), zinc (plasma zinc), folate (plasma folate, erythrocyte folate), vitamin A (total body stores of vitamin A [WRA only], serum retinol, plasma RBP), and vitamin B12 (plasma B12) measured in venous blood samples. Pilot data indicated that vitamin A deficiency was common among children but not WRA in the community; therefore, total body vitamin A stores were selected as the vitamin A outcome for WRA because serum retinol is expected to respond to increased vitamin A intake only when vitamin A stores are low [55]. For lactating women, primary outcomes are concentrations of vitamin A (corrected for milk fat) and vitamin B12 in human milk. Secondary outcomes for women and children include anemia and prevalence of micronutrient deficiencies, malaria and inflammation (CRP and AGP), blood pressure and hypertension, and urinary iodine and sodium. Additional secondary outcomes include dietary intake of bouillon and micronutrients, and morbidity symptoms (all participants); low milk micronutrient concentrations (lactating women); and child development and stool calprotectin (children). **S1 and S2** (**Supplemental Tables 1 and 2**) include detailed lists of primary and secondary outcomes, and **S3 and S3** (**Supplemental Tables 3 and 4**) specify the micronutrient biomarker cutoffs to be used.

#### Data analysis

Statistical analysis plans (SAPs) will be developed and posted publicly prior to conducting any data analysis. For each analysis, a flow diagram for participation will be prepared according to CONSORT guidelines. In most cases, separate analyses will be conducted for each physiological group. Because randomization will occur at the household level and only one individual per physiological group will be enrolled per household, the participants will be considered independent with repeated measurements on the same participant being accounted for with robust standard errors [56]. Additionally, a variable representing the recruitment site of each participant will be included as a control covariate.

The primary analysis will be based on a complete-case intention to treat framework and analysed as a pre-post study design, controlling for baseline measure of the outcome [57]. Testing will be two-sided superiority testing. For continuous outcomes, this will follow an ANCOVA approach with non-normal outcomes transformed prior to analysis (typically log transformed). For binary outcomes, we will use either logistic regression to estimate odds ratios or binomial log-link / modified poisson regression to estimate risk ratios (approach will be pre-specified in each SAP). In addition to the outcome-specific testing, certain analyses may use a multivariate analysis to examine the simultaneous impact of the intervention on multiple outcomes (approach will be pre-specified in each SAP).

In keeping with the randomized design of the study, we will rely on the minimally adjusted analysis (only controlling for baseline measure and recruitment site) as our primary analysis. For selected biomarkers (for example, plasma ferritin), the primary analysis will use inflammation- corrected outcomes and/or a minimally adjusted analysis that includes markers of inflammation (CRP and AGP) at each time point, if biologically relevant and if the outcome is associated with the markers of inflammation in bivariate analyses (details will be pre-specified in each SAP). In a secondary analysis, we will estimate adjusted parameters by including variables that are strongly associated with the outcome to potentially improve the precision of our estimates (i.e., decrease the standard errors). Variable selection approach will be pre-specified in each SAP. Additionally, pre-specified effect modification testing will be conducted by including an interaction term in minimally adjusted models.

Substantial attrition (>20% loss) after randomization will trigger sensitivity analyses to assess the impact of missingness on inference. This will be conducted either by multiple imputation or inverse probability of censoring-weighted analysis (pre-specified in each SAP). We will also conduct a per protocol analysis using data on adherence to identify the subgroup of participants in “high compliance” households.

A qualitative SAP will also be developed for analysis of the OAA focus group data. FGD transcripts will be translated and transcribed to English by two independent parties. A codebook will be developed, and two researchers will independently code the transcripts with Cohen’s Kappa used to assess intercoder reliability (ICR); transcripts with an ICR <0.6 [58] (less than moderate agreement) will trigger discussion and recoding. Selection of qualitative themes will occur through discussions with the greater research team.

#### Sample size

Detection of an effect size of 0.26 SDs was selected based on calculations of the change in total body vitamin A stores among WRA and iron stores among women and children over the 9-mo trial duration; details are presented in the Technical Appendix to the study protocol, available at https://osf.io/t3zrn/). Assuming this effect size and a level of significance of α=0.05 with 80% power we will target an analytic sample size of 234 per physiological group per intervention group. Although this sample size calculation is based on analysis of a single cross-sectional measurement, final models will incorporate a baseline measurement and repeated endline measurements to provide additional statistical power.

Attrition will be accounted for at multiple time points due to the multi-staged study design involving recruitment, screening and baseline study visits prior to randomization to study group. Overall attrition rates were estimated using cross-sectional pilot survey data and are set at ∼32% for children 2-5 y of age and lactating women and at 54% for WRA to account for additional participant exclusion due to pregnancy during the intervention period. Therefore, the recruitment targets will be 690 lactating women, 690 children 2-5 y of age, and 1028 WRA; each split evenly by intervention group.

### Safety and ethics

#### Ethical and institutional approvals

This trial, including the investigational product, has been reviewed and approved by the Ghana Food and Drug Authority (FDA, clinical trial certificate number FDA/CT/2213), the Ghana Health Service Ethical Review Committee (GHS- ERC) (GHSERC ID: 024/11/21), and the UCD, USA, Institutional Review Board (IRB) Human Subjects Committee (IRB ID: 1837253). The trial was registered on ClinicalTrials.gov (NCT05178407) and the Pan-African Clinical Trial Registry (PACTR202206868437931). Any protocol modifications will be submitted to the FDA, GHS-ERC, and UCD IRB for review, and the trial registries will be updated with approved modifications.

We will seek permission from the Regional (Northern Region) and District (Tolon and Kumbungu) Directors of Health of Services as well as community leaders before recruiting participants into the study. With the assistance of the local health personnel, we will then hold general informational meetings with community leaders of each of the communities in the catchment area about the objectives, methods and possible benefits of the study for the study participants and communities.

#### Safety monitoring

A Data and Safety Monitoring Board (DSMB) will be established for this project. The board will consist of 5 members (3 in Ghana, 2 international) not otherwise involved in this study, without competing interests, and who are experts in child health and nutrition research (with one or more members with expertise in medicine, infectious disease, and/or biostatistics). The DSMB will meet at predefined intervals (with intermediate meetings added if deemed necessary by the committee or investigators) to review study progress (including recruitment, loss to follow up, and protocol compliance) and safety monitoring data. The DSMB may recommend actions or modifications, which will be addressed by the study investigators, who will then notify and/or seek approval from the relevant ethical review committees, as appropriate. Blinded interim analysis of severe adverse advents (SAE) and selected adverse events (AE, such as diarrhea and malaria) will be conducted for safety considerations and reviewed by the DSMB. No interim analyses related to intervention efficacy are pre-planned.

The GHS-ERC, Ghana FDA (which regulates clinicals trials in Ghana), UCD IRB, or the DSMB may request unblinded safety data to discuss in closed sessions. Unblinding of the intervention group may be requested if there is a pattern of SAEs which may be related to study treatment where the DSMB or other regulatory agency feels there is “potential for harm”. In this case, the randomization code will be provided by one of the two code keepers.

Suspected SAEs, such as deaths or hospitalization of study participants, will be reported immediately to the principal investigators, who will notify the GHS-ERC, the Ghana FDA, the UCD IRB, and the DSMB in a timely manner as required by their policies; these agencies and committees will then determine whether modifications are required to ensure participant safety and welfare.

To minimize chance of exposure to COVID-19, we have developed Standard Operating Procedures for our field staff and study participants based on recommendations by the Government of Ghana and the GHS-ERC. A physician assistant will refer participants for medical advice and treatment if they are found to have severe malnutrition (children), severe anemia (children or WRA), malaria infection (children or WRA), hypertension (WRA), or other signs of severe illness. Morbidity will be monitored at biweekly home visits and participants will be referred for follow up treatment based on symptoms reported during these visits. The cost of the national health insurance annual premium will be covered for all randomized participants.

Participants who become pregnant will be excluded from the study for scientific reasons (effects of pregnancy on micronutrient biomarker concentrations, and lack of validation of the RID method for pregnant women). Although exclusion of study participants due to pregnancy is not based on any safety concerns, we will follow all index participants who become pregnant until the resolution of their pregnancy (miscarriage, still birth, live birth, etc.) via telephone interview to monitor pregnancy outcomes.

### Trial management

The trial will be implemented by investigators at the University of Ghana and UCD, who jointly developed the trial protocol and all data collection procedures and instruments. Data collection staff will be hired by the University of Ghana, including supervisors, anthropometrists, phlebotomist and laboratory staff, nurse, medical assistant, enumerators, an information technology manager, data entry clerks, and an inventory manager. Staff will participate in intensive training on research ethics, the study protocol, and data collection procedures, including didactic sessions with accompanying quizzes, demonstrations, and pilot-testing. UCD staff will support the study implementation through in-person supervision and remote data monitoring. Investigators from both universities meet weekly to discuss study progress. As the trial is subject to audit at the discretion of the Ghana FDA, the study investigators have contracted with a Local Monitor who is responsible for conducting Good Clinical Practices training of field staff, assessing and approving site readiness to begin data collection, and conducting regular visits to the study site to monitor study implementation. The Regional (Northern Region) and District (Tolon and Kumbungu Districts) Directors of Health were informed about the project on a regular basis throughout pilot research conducted in 2020-2022, and these interactions will continue through trial implementation and results dissemination.

This study is part of a larger initiative supported by the Bill & Melinda Gates Foundation that is focused on generating evidence to inform discussions of bouillon fortification policy among countries in West Africa. Helen Keller International received support from the Bill & Melinda Gates Foundation for research and policy engagement activities related to bouillon fortification in West Africa (INV-007916), that includes supporting the activities of multi-stakeholder bouillon Country Working Groups in Nigeria, Senegal, and Burkina Faso. Helen Keller

International also administers a subaward to UCD for conduct of this trial as well as a portfolio of modelling research focused on estimating the impacts, costs, and cost-effectiveness of micronutrient-fortified bouillon. The larger initiative includes research on technical aspects of bouillon fortification by the Commonwealth Scientific and Industrial Research Organisation (CSIRO) and Research Institutes of Sweden AB (RISE), in collaboration with a consortium of representatives of companies involved in production of bouillon or micronutrient fortificants. These consortium partners developed the non-proprietary unfortified bouillon cube which served as the base formulation for the multiple micronutrient-fortified cubes tested in this trial. All members of this large initiative were given the opportunity to review the study protocol and provide comments. However, the trial investigators selected the micronutrient levels in the investigational bouillon cubes and retain all final decision-making authority related to all aspects of study design, implementation, and dissemination (subject to requirements of the Ghana FDA and ethical committees). Regular updates on study progress will be provided to consortium partners through email and virtual meetings.

### Dissemination

The results from this study will be disseminated in national and international conferences and published as Open Access publications in peer-reviewed journals. Authorship will be determined based on international criteria [59]. Dissemination workshops will be held with national stakeholders in Ghana, and with regional and district stakeholders in the study area. Study results will also be disseminated through the network of bouillon-related partners and stakeholders associated with this project, with particular focus on the West Africa region. Specific dissemination activities will be informed by discussions of the trial Dissemination Advisory Group, convened by University of Ghana, UCD, and Helen Keller International, and consisting of representatives from academia and the public sector in Ghana, Burkina Faso, Senegal, and Nigeria. Publications, Statistical Analysis Plans, and other study resources will be posted on Open Science Framework (https://osf.io/t3zrn/).

### Trial status

Recruitment began on January 16, 2023. Data collection is expected to be completed in mid- 2024.

## Discussion

The burden of micronutrient deficiencies continues to be unacceptably high in many parts of the world. In West Africa, bouillon fortification has the potential to reduce this burden. This study will generate the first information on the effects of micronutrient-fortified bouillon cube on the micronutrient content of human milk and the micronutrient status of women and young children, populations that are vulnerable to micronutrient deficiency. As such, the study serves as “proof of concept” regarding the potential efficacy of micronutrient-fortified bouillon to achieve adequate micronutrient status and reduce the consequences of deficiency. This information will be useful to national, regional and global stakeholders involved in assessing the potential for fortified bouillon cubes to contribute to addressing deficiencies in one or more of the micronutrients examined.

Research and policy discussions related to bouillon fortification must consider the generalizability of study results, with specific attention to the 1) study population diets and other characteristics, 2) micronutrient concentrations in the fortified bouillon, and 3) differences between research and programmatic/commercial contexts for bouillon. The trial is being conducted in a community with frequent bouillon intake and high prevalence of micronutrient deficiency, and the micronutrient concentrations of the cubes tested in this trial were selected as those likely to be both efficacious (i.e., increasing micronutrient status) and safe (i.e. not causing adverse effects) in this context. While these characteristics also apply to many communities throughout West Africa, there is great variation within and across countries in bouillon consumption and micronutrient deficiency [14, 16]. Discussions on bouillon fortification standards must consider the impacts on population subgroups with both adequate and inadequate micronutrient intake, and populations with varying bouillon consumption.

Where countries move ahead with bouillon fortification standards (or at the regional level, as cooking oil and wheat flour fortification are implemented in West Africa), decisions around selection of the desired micronutrient concentrations in the standards should use population-specific data on dietary intake as a starting point [29] rather than the levels tested in this trial. Micronutrient concentrations in standards may be different than those tested in this study, and standards may also include micronutrients that were not tested in this study. Modelling of dietary intake or household consumption surveys data can help inform what micronutrients and what concentrations will meet dietary gaps for target population subgroups while minimizing intakes above the UL [60]. Standards must also consider technical and commercial constraints, such as required changes to product formulas and manufacturing processes, premix cost, and the ability to pass along some or all of these costs to bouillon consumers. Moreover, it is important to note that in the trial context we are maintaining bouillon cubes in climate-controlled storage conditions (up to the point of delivery to the household) to ensure acceptability and micronutrient stability of the cubes; these characteristics will have to be confirmed under commercial conditions.

In this trial, bouillon is regularly delivered to participant households, free of charge. The household bouillon ration is capped to avoid extra-household sharing and artificially inflating the impact of fortified bouillon if bouillon consumption were to increase due to providing bouillon free of charge to participating households. Nevertheless, in a program setting the effects of fortified bouillon may be diluted if less-expensive, unfortified bouillon products are available or if households reduce their bouillon consumption in response to price increases due to fortification. Moreover, program compliance with standards is a challenge with other types of large-scale food fortification programs [11] and especially for easily imported products, therefore highlighting the need for effective monitoring and evaluation programs to secure the effectiveness of any future bouillon fortification programs.

Given the wide and equitable reach of bouillon in West Africa, including among populations at risk of micronutrient deficiencies who do not consume other fortified foods, addition of micronutrients to bouillon holds great potential to reduce deficiencies. This study will provide evidence regarding the efficacy of bouillon fortification in a specific West African context for improving micronutrient status. This new evidence will inform ongoing discussions on large-scale food/condiment fortification and other strategies to reduce the burden of micronutrient deficiencies and their consequences in the region.

## Data Availability

Consistent with policy of the Bill & Melinda Gates Foundation, data and forms will be made available online at osf.io/t3zrn/ three years after the completion of data collection.

## Acknowledgments

We gratefully acknowledge the contributions of the local medical monitor Dr. Mohammed Hafiz Kanamu (Tamale Teaching Hospital) and the local study implementation monitors Michael Ntiri and Morrison Asiamah. We thank Dr. Bess Caswell (USDA Western Human Nutrition Research Center) and Boateng Bannerman (University of Ghana, Legon) for assistance with study blinding and Professor Sam Newton (Kwame Nkrumah University of Science and Technology (KNUST)), Dr. Justice Moses K. Aheto (University of Ghana, Legon), Dr. Edward Frongillo (University of South Carolina), Dr. Agartha Ohemeng (University of Ghana, Legon), and Prof Andrew Prendergast (Queen Mary University of London) for serving on the Data and Safety Monitoring Board.

We thank GB foods for producing the trial-specific micronutrient-fortified bouillon cubes for this study, particularly Félix Sancho, Juan Carlos Velasco, and Patricia Lopez. We thank DSM for preparing the micronutrient premix and Symrise for preparing the specific flavouring used in the trial cubes. Colleagues at the Commonwealth Scientific and Industrial Research Organisation (CSIRO), along with members of a consortium created the model bouillon formulation used in this study and advised on technical issues related to bouillon storage and management.

The Bill & Melinda Gates Foundation Design Analyze Communicate (DAC) team provided helpful comments on an early version of the study protocol and facilitated trial design simulations by Mediana. We further appreciate the technical and overall guidance of Dipika Matthias (PATH), Ann Tarini (independent consultant), and colleagues at Helen Keller International. We acknowledge the UCSF MLK Cores Research Facility for support with sample analysis plans. Finally, we thank Yakubu Adams and Felix Kyereh (University of Ghana) and Dr. Lindsay Allen (USDA) for contributions to study preparations, Dr. Elizabeth Prado (UC Davis) for guidance on child development assessments, Lauren Thompson (UC Davis) for assistance with stool collection protocols, Dr. Michael Green (The Pennsylvania State University), Dr. Anthony Oxley and Dr. Alice Goddard (Newcastle University) for support with RID assessment.

---

**S1.** Table of primary outcomes.

S1. Table of primary outcomes
| Physiological group | Primary outcomes |
| --- | --- |
| Non-pregnant, non-lactating women of reproductive age (WRA) | • Change in micronutrient status after 38 weeks:<br>○ Total body vitamin A stores and liver vitamin A concentration (measured by retinol isotope dilution)<br>○ Plasma ferritin and soluble transferrin receptor (sTfR) concentrations, and calculated body iron stores (BIS)<br>○ Plasma zinc concentration<br>○ Erythrocyte folate concentrations<br>○ Plasma vitamin B12 concentration<br>• Change in hemoglobin (Hb) concentration after 38 weeks |
| Children 2-5 y | • Change in micronutrient status after 38 weeks:<br>○ Plasma retinol binding protein (RBP) concentrations<br>○ Plasma ferritin and soluble transferrin receptor (sTfR) concentrations, and calculated body iron stores (BIS)<br>○ Plasma zinc concentration<br>○ Plasma folate concentration<br>○ Plasma vitamin B12 concentration<br>• Change in hemoglobin concentration after 38 weeks |
| Non-pregnant, lactating women | • Change in breast milk vitamin A concentration after 12 weeks (controlling for milk fat content)<br>• Change in breast milk vitamin B12 concentration after 12 weeks |

**S2.** Table of secondary outcomes.

S2. Table of secondary outcomes
| Target group | Secondary outcomes |
| --- | --- |
| Household | Household consumption of bouillon (total and per capita)<br>Adherence (consumption of study-provided bouillon, expressed as % of total household bouillon consumption) |
| Non-pregnant, non-lactating women of reproductive age | Change in serum or plasma retinol and retinol binding protein (RBP) concentrations after 38 weeks<br>Change in serum folate concentrations after 38 weeks<br>Change in urinary iodine concentrations after 38 weeks<br>Urinary sodium concentrations, and sodium:potassium ratio<br>Change in prevalence of micronutrient deficiency after 38 weeks<br>Change in prevalence of anaemia after 38 weeks<br>Inflammation (concentrations of C-reactive protein (CRP) and alpha-1-acid glycoprotein (AGP) and prevalence of elevated values, and other markers including amyloid A, monocyte chemoattractant protein-1 (MCP-1), IL-6, IL-10, IL-1beta, and adiponectin at baseline and endline)<br>Current or recent malaria (by rapid diagnostic test; RDT) at baseline and endline<br>Morbidity (cumulative days of symptoms over study duration)<br>Dietary intake of bouillon, salt, fortifiable foods, energy, macronutrients, and micronutrients<br>Blood pressure and hypertension prevalence |
| Children 2-5 y | Change in urinary iodine concentrations after 38 weeks<br>Change in prevalence of micronutrient deficiency after 38 weeks<br>Change in prevalence of anaemia after 38 weeks<br>Change in plasma retinol concentrations after 38 weeks<br>Inflammation (concentrations of C-reactive protein (CRP) and alpha-1-acid glycoprotein (AGP) and prevalence of elevated values) at baseline and endline<br>Current or recent malaria (by RDT) at baseline and endline<br>Faecal calprotectin concentrations at baseline and endline<br>Faecal microbiota at baseline and endline<br>Morbidity (cumulative days of symptoms over study duration)<br>Dietary intake of bouillon, salt, fortifiable foods, energy, macronutrients, and micronutrients<br>Anthropometric measurements (change in height-for-age and weight-for-height Z-scores; prevalence of stunting and wasting)<br>Change in Malawi Development Assessment Tool (MDAT) scores in 4 domains (gross motor, fine motor, language, social) after 38 weeks<br>Change in Early Years Toolbox (EYT) scores after 38 weeks |
| Non-pregnant, lactating women | Change in prevalence of low milk nutrient concentrations after 12 weeks<br>Dietary intake of bouillon, salt, fortifiable foods, energy, macronutrients, and micronutrients<br>Morbidity (cumulative days of symptoms over study duration) |

**S3.** Table of specification and definitions of hemoglobin and micronutrient biomarker variables for children 2-5 years of age^1^.

| Outcome variables | Definitions of dichotomous outcome variables |
| --- | --- |
| Hemoglobin (Hb; g/L) |  |
| Anemia | Hb < 110 g/L (WHO, 2011) |
| Mild anemia | Hb 100-109 g/L |
| Moderate to severe anemia | Hb 70-99 g/L |
| Severe anemia | Hb < 70 g/L |
| Plasma ferritin (µg/L) |  |
| Iron deficiency (low plasma ferritin concentration) | Plasma ferritin < 12 µg/L (WHO, 2011) |
| Plasma soluble transferrin receptor concentration (sTfR; mg/L) |  |
| Elevated plasma sTfR concentration | Plasma sTfR > 8.3 mg/L (Erhardt, Estes, Pfeiffer, Biesalski, & Craft, 2004) |
| Any iron deficiency | Plasma ferritin < 12 µg/L and/or sTfR > 8.3 mg/L |
| Iron deficiency anemia | (plasma ferritin < 12 µg/L and/or sTfR > 8.3 mg/L) and Hb < 110 g/L (World Health Organization) |
| Low ferritin and anemia | Plasma ferritin < 12 µg/L and Hb < 110 g/L (World Health Organization) |
| Body iron stores (BIS; mg/kg) |  |
| Low BIS | < 0 mg/kg (Cook, Flowers, & Skikne, 2003) |
| Plasma zinc concentration (µg/dL) |  |
| Low plasma zinc concentration | Plasma zinc < 65 µg/dL (Brown et al., 2004) |
| Plasma retinol concentration (µmol/L) |  |
| Low retinol | Plasma retinol < 0.70 µmol/L (WHO, 2011) |
| Marginal retinol | Plasma retinol < 1.05 µmol/L |
| Plasma retinol binding protein concentration (RBP; µmol/L)<sup>2</sup> |  |
| Low RBP | Study-specific cut-off to be determined |
| Marginal RBP | Study-specific cut-off to be determined |
| Plasma folate concentrations (nmol/L) |  |
| Low folate | < 10 nmol/L (L. B. Bailey et al., 2015) |
| Low folate and anemia | Plasma folate < 10nmol/L and anemia (Hb < 120 g/L) |
| Plasma vitamin B12 concentrations (pmol/L) |  |
| Low vitamin B12 | < 148 pmol/L (Allen et al., 2018) |
| Depleted vitamin B12 | < 221 pmol/L |
| Low B12 and anemia | Vitamin B12 < 148 pmol/L and anemia (Hb < 120 g/L) |

**S4.**
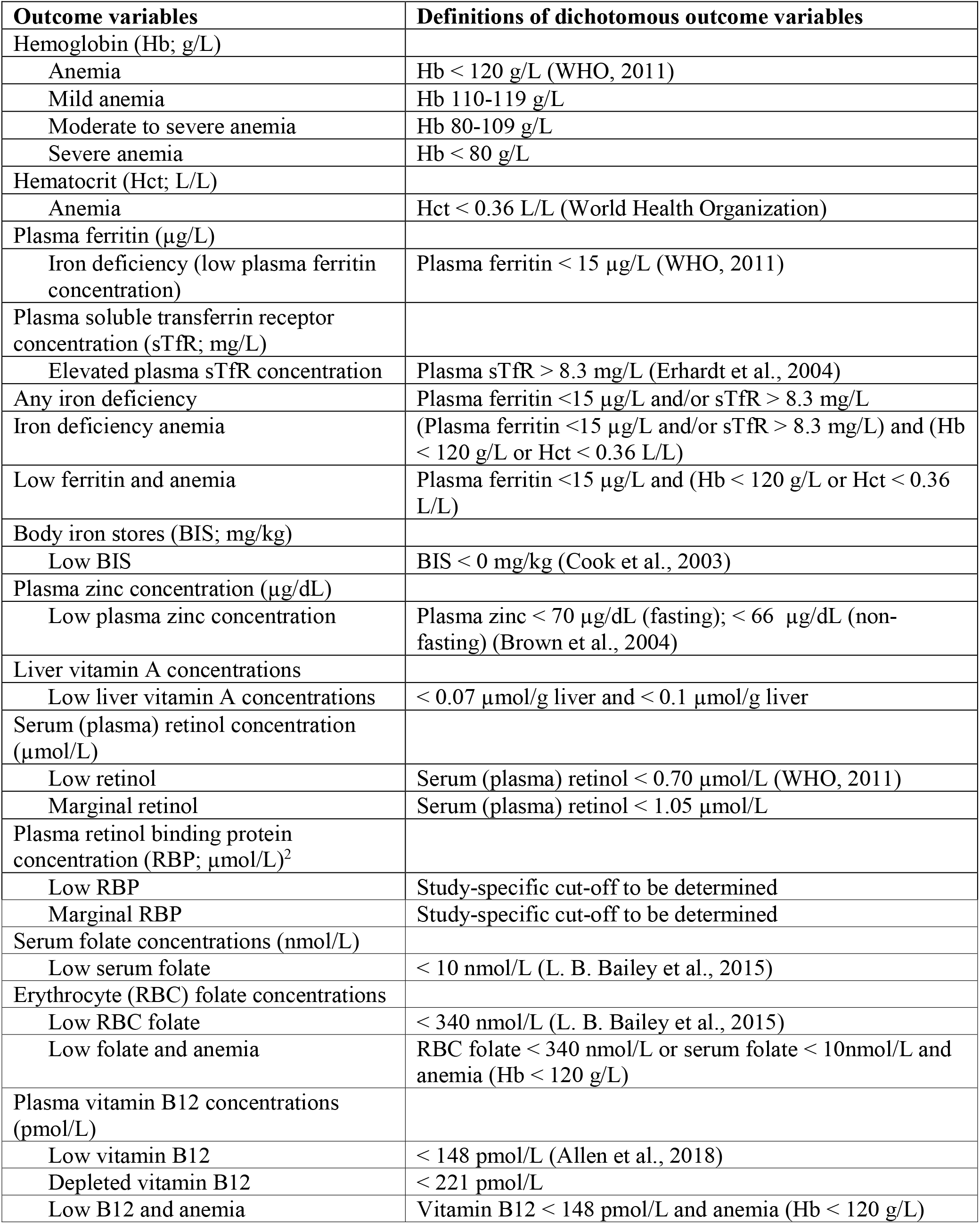
Table of specification and definitions of hemoglobin and micronutrient biomarkers variables for non-pregnant, non-lactating women of reproductive age^1^

## S5. Supplemental Methods: Inclusion, Exclusion, and Deferral Criteria

### Inclusion/exclusion criteria at recruitment (home visit)

#### Household

Inclusion criteria are:

i. Head of household provides oral consent for the participation of household members (index participants), and willingness to have study-provided bouillon cubes used in their household cooking for the next 10 months;

Exclusion criteria are:

i. Reported chronic medical condition requiring frequent blood transfusion (e.g. severe forms of thalassemia) among any household members;
ii. Current participation of any household member in a clinical trial; (iii)Reported shrimp, wheat, milk, soy, eggs, celery, fish, or mollusk allergy, or a previous adverse reaction to bouillon by the participant or any member of their household.

#### Non-pregnant, non-lactating women of reproductive age (WRA)

Inclusion criteria are:

i. Non-pregnant non-lactating women of reproductive age (15 - 49 years);
ii. Signed the informed consent form (or in the case of adolescents 15-17 years of age, unmarried and still living with their parents, assent provided from the index participant and consent from a parent or guardian);
iii. Planning to remain in the study area for the next 10 months;
iv. Willing to use study-provided bouillon in household cooking for the next 10 months;
v. Not planning to become pregnant during the next 10 months.

Potential participants will be excluded if any of the following apply:

i. Severe illness warranting immediate hospital referral;
ii. COVID-19 diagnosis or exposure^1^ in the previous two weeks *[individual may repeat eligibility assessment once after a deferral period of at least 2 weeks];*
iii. Presence of morbidity symptoms suggesting COVID-19 infection (fever [temperature ≥ 38°C], chills/shaking, dry cough, shortness of breath or difficulty breathing, loss of smell or taste within the past 72 hours) *[individual may repeat eligibility assessment once after a deferral period of at least 2 weeks];*
iv. Chronic severe medical condition (e.g. malignancy) or congenital anomalies requiring frequent medical attention or potentially interfering with nutritional status;
v. Unable to provide informed consent due to impaired decision-making abilities.

#### Children 2-5 years of age (24-59 mo)

Inclusion criteria are:

i. Child 2-5 years of age (24-59 mo);
ii. Signed informed consent for the child’s participation from a parent or guardian;
iii. Planning to remain in the study area for the next 10 months;
iv. Caregiver willing to use study-provided bouillon in household cooking for the next 10 months.

Potential participants will be excluded if any of the following apply:

i. Severe illness warranting immediate hospital referral;
ii. COVID-19 diagnosis or exposure in the previous two weeks *[individual may repeat eligibility assessment once after a deferral period of at least 2 weeks];*
iii. Presence of morbidity symptoms suggesting COVID-19 infection (fever, [temperature ≥ 38°C], chills/shaking, dry cough, shortness of breath or difficulty breathing, loss of smell or taste within the past 72 hours) *[individual may repeat eligibility assessment once after a deferral period of at least 2 weeks];*
iv. Mid-upper arm circumference (MUAC) < 11.5 cm;
v. Chronic severe medical condition (e.g. malignancy) or congenital anomalies requiring frequent medical attention or potentially interfering with nutritional status.

#### Lactating women

Inclusion criteria are:

i. Non-pregnant women of reproductive age (15 - 49 years); currently breastfeeding a child who is 4-18 months of age;
ii. Signed the informed consent form (or in the case of adolescents 15-17 years of age, unmarried and still living with their parents, provide assent from the index participant and consent from a parent or guardian);
iii. Planning to remain in the study area for the next 4 months;
iv. Planning to breastfeed for the next 4 months;
v. Willing to use study-provided bouillon in household cooking for the next 4 months;
vi. Not planning to become pregnant during the next 4 months.

Potential participants will be excluded if any of the following apply:

i. Pregnancy (determined by self-report);
ii. Severe illness warranting immediate hospital referral;
iii. COVID-19 diagnosis or exposure in the previous two weeks *[individual may repeat eligibility assessment once after a deferral period of at least 2 weeks];*
iv. Presence of morbidity symptoms suggesting COVID-19 infection (fever[temperature ≥ 38°C], chills/shaking, dry cough, shortness of breath or difficulty breathing, loss of smell or taste within the past 72 hours) *[individual may repeat eligibility assessment once after a deferral period of at least 2 weeks];*
v. Chronic severe medical condition (e.g. malignancy) or congenital anomalies requiring frequent medical attention or potentially interfering with nutritional status;
vi. Unable to provide informed consent due to impaired decision-making abilities.

### Exclusion criteria at <u>baseline</u> screening visit (WRA only)

#### Non-pregnant, non-lactating women of reproductive age

Potential participants will be excluded if any of the following apply:

i. Hemoglobin < 80 g/L **at baseline screening visit**;
ii. Severe illness warranting immediate hospital referral;
iii. COVID-19 diagnosis or exposure in the previous two weeks *[individual may repeat eligibility assessment once after a deferral period of at least 2 weeks];*
iv. Presence of morbidity symptoms suggesting COVID-19 infection (fever [temperature ≥ 38°C], chills/shaking, dry cough, shortness of breath or difficulty breathing, loss of smell or taste within the past 72 hours) *[individual may repeat eligibility assessment once after a deferral period of at least 2 weeks];*
v. Recent diarrhea [≥3 liquid or semiliquid stools in 72 hours]) *[individual may repeat eligibility assessment once after a deferral period of at least 2 weeks];*
vi. Reported consumption of vitamin A-rich foods (e.g., liver) in the previous 24 hours *[individual may repeat eligibility assessment once after a deferral period of at least 2 weeks];*
vii. Pregnancy (as ascertained via urine pregnancy test for human chorionic gonadotropin, HCG, on the day of isotope dosing);
viii. Incomplete consumption of vitamin A isotope dose;
ix. Positive malaria RDT on the day of isotope dosing *[individual may repeat eligibility assessment once after a deferral period of at least 2 weeks];*
x. CRP > 5 mg/L on the day of isotope dosing *[individual may repeat eligibility assessment once after a deferral period of at least 2 weeks]*.

### Exclusion criteria at baseline visit

#### Non-pregnant, non-lactating women of reproductive age

i. Hemoglobin < 80 g/L;
ii. Severe illness warranting immediate hospital referral;
iii. COVID-19 diagnosis or exposure in the previous two weeks *[individual may repeat eligibility assessment once after a deferral period of at least 2 weeks];*
iv. Presence of morbidity symptoms suggesting COVID-19 infection (fever [temperature ≥ 38°C], chills/shaking, dry cough, shortness of breath or difficulty breathing, loss of smell or taste within the past 72 hours) *[individual may repeat eligibility assessment once after a deferral period of at least 2 weeks];*
v. Pregnancy (determined by self-report).

#### Children 2-5 years of age

i. Hemoglobin < 70 g/L;
ii. Severe illness warranting immediate hospital referral;
iii. COVID-19 diagnosis or exposure in the previous two weeks *[individual may repeat eligibility assessment once after a deferral period of at least 2 weeks];*
iv. Presence of morbidity symptoms suggesting COVID-19 infection (fever [temperature ≥ 38°C], chills/shaking, dry cough, shortness of breath or difficulty breathing, loss of smell or taste within the past 72 hours) *[individual may repeat eligibility assessment once after a deferral period of at least 2 weeks];*
v. Severe acute malnutrition at baseline (weight-for-height Z-score < -3 SD or bilateral oedema).

#### Lactating women

i. Severe illness warranting immediate hospital referral; (xi) COVID-19 diagnosis or exposure in the previous two weeks *[individual may repeat eligibility assessment once after a deferral period of at least 2 weeks];* (xii) Presence of morbidity symptoms suggesting COVID-19 infection (fever [temperature ≥ 38°C], chills/shaking, dry cough, shortness of breath or difficulty breathing, or loss of taste or smell within the past 72 hours) *[individual may repeat eligibility assessment once after a deferral period of at least 2 weeks];*
ii. (ii) Pregnancy (determined by self-report);
iii. (iii) Cessation of lactation, or planning to discontinue breastfeeding in the next three months.

### Exclusion criteria during course of the intervention

#### Non-pregnant, non-lactating women of reproductive age

i. Pregnancy (determined by self-report);
ii. Pregnancy (as ascertained via urine pregnancy test for human chorionic gonadotropin at the endline 1 visit [pre-endline isotope dosing]).

#### Lactating women

i. Cessation of lactation (determined by self-report);
ii. Pregnancy (determined by self-report).

**Note:** If a COVID-19 exposure or diagnosis, or if COVID-19-related morbidity symptoms are reported during biweekly visits during the intervention trial, the participant will be allowed to remain enrolled, but field staff will follow procedures for “no contact” study activities (e.g., “no contact” bouillon ration drop-off) for a 2-week period.

### Deferral criteria for endline assessments

Replicate endline assessments (3 weeks apart for blood biomarkers and 1 week apart for breast milk biomarkers) will be conducted for each of the primary outcomes except estimation of total body vitamin A stores among women. For all participants, each endline assessment will be deferred if any of the following apply:

i. COVID-19 exposure or positive test in the previous two weeks *[individual may repeat eligibility assessment once after a deferral period of at least 3 weeks for WRA and children or at least 2 weeks for lactating women];*
ii. Presence of morbidity symptoms suggesting COVID-19 infection (fever [temperature ≥ 38°C], chills/shaking, dry cough, shortness of breath or difficulty breathing, or loss of taste or smell within the past 72 hours) *[individual may repeat eligibility assessment once after a deferral period of at least 3 weeks for WRA and children or at least 2 weeks for lactating women]*.

For a participant who has both endline visits deferred, the maximum duration of participation in the study would be 44 weeks (that is, if Endline 1 is deferred from 35 weeks to 38 weeks and Endline 2 is deferred from 41 weeks to 44 weeks). In this case the household would continue to receive their usual supply of study bouillon cubes until the participants complete the Endline 2 visit. Based on pilot data we expect this situation to be rare.

Note: If a non-pregnant, non-lactating WRA has a hemoglobin concentration < 80 g/L, or a 2-5 year old child has a hemoglobin concentration < 70 g/L or a WHZ < -3 SD at the first endline visit, participants may remain in the trial, but will not participate in the second endline blood sample.

**Additional** exclusion criteria for pre-endline isotope dose (WRA only):

#### Non-pregnant, non-lactating women of reproductive age

The pre-endline vitamin A isotope dose will be administered after the endline 1 blood sample. The isotope dose will not be given if any of the following apply:

i. Hemoglobin < 80 g/L

**Additional** deferral criteria for pre-endline isotope dose (WRA only):

#### Non-pregnant, non-lactating women of reproductive age

The pre-endline vitamin A isotope dose will be administered after the endline 1 blood sample. The isotope dose will be deferred if any of the following apply:

i. Recent diarrhea [≥3 liquid or semiliquid stools in 72 hours]) *[individual may repeat eligibility assessment once after a deferral period of at least 3 weeks];*
ii. Reported consumption of vitamin A-rich foods (e.g., liver) in the previous 24 hours *[individual may repeat eligibility assessment up once after a deferral period of at least 3 weeks];*
iii. Positive malaria RDT on the day of isotope dosing *[individual may repeat eligibility assessment once after a deferral period of at least 3 weeks];*
iv. CRP > 5 mg/L on the day of isotope dosing *[individual may repeat eligibility assessment once after a deferral period of at least 3 weeks]*.

## Footnotes

1 Exposure based on the WHO definition: “A contact is defined as anyone who had direct contact or was within 1 metre for at least 15 minutes with a person infected with the virus that causes COVID-19.” https://www.who.int/news-room/q-a-detail/coronavirus-disease-covid-19-contact-tracing. Diagnosis may be based on positive test or diagnosis by a medical professional based on clinical symptoms.

## Notes

### Competing Interest Statement

The authors have declared no competing interest.

### Clinical Trial

NCT05178407

### Funding Statement

This work was supported, in whole or in part, by a grant from Helen Keller International (66504-UCD-01), through support from the Bill & Melinda Gates Foundation (INV-007916), to the University of California, Davis.

### Author Declarations

The Ghana Food and Drug Authority (FDA) gave approval for this work (clinical trial certificate number FDA/CT/2213).

The Ghana Health Service Ethical Review Committee (GHS- ERC) of the Ghana Health Services gave ethical approval for this work (GHSERC ID: 024/11/21).

The Institutional Review Board (IRB) Human Subjects Committee of the University of California, Davis, gave ethical approval for this work (IRB ID: 1837253).

